# Diagnostic Performance of GeneXpert HCV VL Finger Stick and GeneXpert HCV Viral Load Compared to RT-PCR in POCT settings : Establishing the Diagnosis of Chronic Hepatitis C in Hemodialysis Patients

**DOI:** 10.1101/2025.05.02.25326788

**Authors:** Andri Sanityoso Sulaiman, Pringgodigdo Nugoroho, Darlene Raudhatul Bahri, Desti Rachmani

## Abstract

**Background:** The most widely utilized method for detecting HCV RNA is Nucleic Acid Testing (NAT) via RT PCR. In response to the need for more accessible and faster testing, particularly in high risk populations, an accessible method : Point of Care Molecular Testing (PoCT). International studies have demonstrated the use of fingerstick samples with this PoCT molecular method among injecting drug users in prisons, where HCV RNA PoCT exhibited a sensitivity of 98.4% for detecting viral loads (>4 IU/mL), and 100% sensitivity for quantifiable viral loads (≥10, 1000, and 3000 IU/mL), with a specificity of 100%. This PoCT testing is also a key element in the WHO’s strategy for achieving the Hepatitis elimination target by 2030. However, its application in Indonesia remains limited, and as such, the efficacy of this simple testing approach has yet to be fully evaluated in comparison to real-time PCR.

**Method:** This study is a diagnostic test with a cross-sectional design. There were 57 CKD on Hd patients in Cipto Mangunkusumo and Pelni Hospital tested by GeneXpert Fingerstick, Plasma and confimerd by PCR Cobas Taqman

**Result:** The probability test comparing the Fingerstick, GeneXpert, and Cobas Taqman 96 platforms yielded a p-value of <0.001, indicating a highly significant correlation. Both the Fingerstick and GeneXpert HCV viral load assays demonstrated exceptional diagnostic performance, achieving 100% sensitivity, specificity, negative predictive value (NPV), and positive predictive value (PPV)

**Conclusion:** The GeneXpert fingerstick POCT may serve as a simple and reliable point-of-care method for detecting hepatitis C in patients with chronic kidney disease undergoing hemodialysis

## Introduction

Hepatitis C-related liver cancer (LCDHC) is a leading cause of cancer-related death globally. In 2020, liver cancer was the most common malignant solid tumor in the globe, accounting for over 905,700 new cases and 830,200 reported deaths.. [1]. Based on GLOBOCAN 2020 data, liver cancer ranked as the third most common cause of cancer-related deaths and the sixth most commonly diagnosed cancer globally. [2]. In addition, there were about 140000 deaths due to liver cancer related to hepatitis C in 2019 (1), Yang Jianqin et all described complete epidemiological data on LCDHC, including incidence, mortality, and disability-adjusted life years (DALYs), at the regional, national, and global levels is still unavailable. Yang Jianqin and colleagues documented the burden of LCDHC in China between 1999 and 2019. Worldwide, liver cancer was responsible for 12,528,422 (11,400,671–13,687,675) DALYs, 534,364 (486,550–588,639) incident cases, and 484,577 (95% UI 444,091–525,798) deaths in 2019. From 1990 to 2019 the numbers increased This highlight need for enhanced understanding and the allocation of resources for effective disease management and prevention(2,3) Hepatitis C virus (HCV) associated diseases are rarely diagnosed solely on the basis of clinical symptoms, as the infection often remains asymptomatic or exhibits only mild symptoms over prolonged periods. As a result, diagnosis is typically established following the incidental identification of laboratory markers indicative of HCV infection. (4) Early diagnosis and prompt initiation of treatment are essential to avert the progression to severe liver complications and to mitigate the risk of continued transmission. Hepatitis C in patients on dialysis needs accurate diagnosis and follow-up for realistic medical management. Serological tests, such as enzyme-linked immunosorbent assay, are commonly used for screening, but they may have lower sensitivity in dialysis patients due to the reduced immune response. Molecular tests, such as HCV RNA quantification, are preferred for diagnosis and monitoring of treatment response. (5) In particular, dialysis patients are at a significantly higher risk of HCV infection compared to the general population, with the disease tending to occur more frequently and severely in this group. (5,6) Meta-analysis in 2020 showed in hemodialysis patients have a higher risk of contracting HCV infection due to factors such as shared dialysis equipment, improper infection control practices, and the need for frequent blood draws. Early diagnosis and management of HCV infection are crucial in this population to prevent complications, such as cirrhosis and HCC.(7) Point-of-care testing has gained increasing attention as a means to improve diagnosic and management of HCV infection in hemodialysis patients. These tests, which can be performed at the bedside or in the clinic, offer several advantages over conventional laboratory-based testing, including faster turnaround times, easier sample collection, and the potential for increased patient engagement and adherence to treatment. The GeneXpert Plasma test, a rapid, fully-automated molecular assay that detects and quantifies HCV RNA directly from plasma samples, has been reported to be a sensitive and specific molecular test for HCV. This test offers the benefit of providing a measurable result, which can be useful for monitoring disease progression and evaluating how well the treatment is working, although the information may have limited clinical significance.. The Xpert HCV VL Fingerstick assay is a newly developed, CE-marked point-of-care test (POCT) for HCV-RNA that enables sameday clinical decision making. It allows for on the spot diagnosis of active HCV infection, eliminating the need for patients to return for a follow-up visit. (8) The GeneXpert test is a rapid automated molecular assay capable of HCV RNA detection and quantification directly from plasma samples. The advantage of this test is that numerical value is obtained, which is effective in assessing disease progression and response to treatment. The GeneXpert HCV Viral Load Fingerstick assay represents a promising solution to overcome the existing challenges in HCV diagnosis.(9,10) The included studies Umumararungu et al., 2017 and Tang et al., 2022 study examined the application of the GeneXpert HCV RNA Assay among diverse populations, including those receiving care at a military hospital in Rwanda and individuals in point-of-care environments such as HCV clinics and substance use treatment facilities. The diagnostic accuracy of the GeneExpert assay was assessed by comparing its results to those obtained from established laboratory-based HCV RNA detection methods, such as the Abbott RealTime HCV Assay.(11) Several studies have demonstrated the high sensitivity and specificity of the GeneExpert HCV RNA Assay in detecting HCV RNA in both venous blood plasma and fingerstick sample. (12) When tested with plasma samples, one study found the GeneXpert assay to have a sensitivity of 98.4% and a specificity of 99.1%. Another study using finger-stick blood samples reported a sensitivity of 95.7% and a specificity of 100%. The assay’s ability to provide rapid and accurate results, as well as its suitability for use in resource-limited settings, have been highlighted in the literature. (13,14)

A previous study by Iwatomo et al. (2018) compared the performance of GeneXpert® (Cepheid) with Cobas® TaqMan® (Roche) for HCV RNA quantification in a patient population in Cambodia where genotypes 1 and 6 are common. The results showed that 454 patients (77%) had detectable and quantifiable viral loads using the Roche assay, and 195 of them (43%) were identified as having genotype 6 (GT6). GeneXpert® showed 100% sensitivity (95% CI: 99.2– 100.0) and 98.5% specificity (95% CI: 94.8–99.8) in comparison to Roche. GeneXpert® produced same-day results (0-day TAT) with only 1% error, whereas Roche had a 4-day TAT. (23)

## Method

### Design Study

This study is a diagnostic test with a cross-sectional design aimed at evaluate the diagnostic performance of the GeneXpert HCV VL Finger Stick POCT for the quantitative detection of HCV RNA in a specific population of Chronic Kidney Disease (CKD) patients undergoing hemodialysis with positive anti-HCV status. The GeneXpert HCV RNA fingerstick will be compared with GeneXpert HCV Viral Load (Venous blood sample and real-time PCR Cobas Taqman as the reference gold standard. The GXHCV-VL-CE-10 assay was developed by Cepheid. For comparison, samples from the same plasma specimen were also tested using the Abbott RealTime HCV Viral Load assay. The following day, 1 mL of plasma was processed with the GeneXpert device, testing was carried out using the Xpert HCV Viral Load cartridge (GXHCV-VL-CE-10; Cepheid, Sunnyvale). All procedures were performed on a clinic-based 6-color, 4-module GeneXpert R2 system (GXIV-4-L System, 900-0513) with the GeneXpert Dx v4.6a software. (15)

### Patients

57 chronic kidney disease patients on hemodialysis were selected from Cipto Mangunkusumo National and Pelni Hospital in Jakarta. Baseline information, including participant’s age, gender, and HCV antibody status. The study received approval from the Faculty of Medicine Universitas Indonesiia’s Ethics Commite (KET-779/UN2.F1/ETIK/PPM.00/02/2022).

### GeneXpert HCV VL Fingerstick

A small volume of capillary blood collected from the Fingertip, A total of 100 μl of capillary whole blood was obtained using the BD Microtainer™ Contact-Activated Lancet. The blood sample was then collected into a 100 μl EDTA-coated minivette (Sarstedt Minivette POCT).BD MicrotainerTM Contact-Activated Lancet took 100 μl of capillary whole blood and placed it in an EDTA minivette (Sarstedt Minivette POCT 100 μl).. The sample was then immediately loaded into a GeneXpert HCV VL FS cartridge (GX-HCV-FS-CE-10; Cepheid), which is designed for use with 100 µl capillary whole blood (with a limit of quantification at 100 IU/mL and a limit of detection at 40 IU/mL). Results were available within 60 minutes using the GeneXpert system. These rapid tests are simple to use and require only minimal training to perform.

### GeneXpert HCV Viral Load *(Venous blood sample)*

A 1mL of serum or plasma was taken from a venous blood sample, centrifuged, and then placed into the “GXHCV-VL-CE-10” HCV Viral Load cartridge. The entire analysis process took 105 minutes. For point-of-care testing (POCT), both assays were used. The GeneXpert system consists of four modules, which are connected to a laptop and a portable device. The GeneXpert® device and software were set up, patient details were entered, and the Xpert® cartridge was scanned before capillary blood was collected. Once the capillary blood was drawn, it was immediately placed into the cartridge, and the system began analyzing the sample. Thus, the time from puncture to test was less than one minute because each of the four modules operates independently, tests could begin even while the first module was still running, ensuring efficient workflow.

### Reference Gold Standard PCR Cobas® TaqMan

HCV RNA Detection by RT-PCR COBAS AmpliPrep®/COBAS TaqMan® HCV (Roche®, Ref. Nr. 05532264) 750 ul of plasma samples were tested by the Abbott Real Time HCV assay on the automated Abbot m2000sp/m2000rt platform, as per the manufacturer’s instructions. The results from routine testing were documented and served as the basis fot comparative anaysisis in this study. The assay’s lower limit of detection (LOD) and lower limit quantifications (LLOQ) were founf to be comparable. Plasma from venapuncture are centrifuged within 6 hours and stored at −20° then processed with the Cobas® Taqman® analyzer. The Cobas® AmpliPrep / Cobas® TaqMan® HCV test involves three main steps :

1. Specimen preparation: The first step is isolating the HCV RNA from the sample.
2. Reverse transcription: In this step, the isolated RNA is converted into complementary DNA (cDNA) through reverse transcription.
3. PCR amplification and detection: The final step involves amplifying the cDNA through PCR, followed by detection using a dual-labeled oligonucleotide probe that is specific to the target.

The three main steps were specifically designed to bing into a sequence located in the higlu conserved 5’unstranslated region of the HCV genome. Their nucleotide sequences have been carefully optimized to allow for reliable amplification across all HCV genotypes.

## STATISTICAL ANALYSIS

The data for the GeneXpert HCV VL Fingerstick and GeneXpert HCV Viral Load tests were analyzed independently. Sensitivity (the number of HCV RNA positive samples with a positive capillary test / total number of HCV RNA positive samples), specificity (the number of HCV RNA negative samples with a negative capillary test / total number of HCV RNA negative samples), the proportion of individuals with positive capillary test results who actually were HCV RNA positive (positive predictive value [PPV]), the percentage of individuals with negative capillary test results who truly were negative by HCV RNA (negative predictive value [NPV]), and/or the percentage of test results that concordantly (positive or negative) were qualitative were each evaluated. Only the qualitative outcomes (detectable, non-detectable) were included for the sensitivity and specificity analysis. Samples that produced quantifiable results from both methods were analyzed for correlation, with the squared correlation coefficient (R^2^) and the corresponding regression line calculated.

## RESULT

### Baseline Characteristic of Study Participants

A total of 57 patients enrolled between Desember 2022 and July 2024, each had a fingerstick capillary blood, plasma sample and PCR sample. In this study, with a mean age of 52.88 ± 14.04 years. Among them, 66.7% were male, and 63.2% tested positive for HCV antibodies (Table 1.)

**Table 1.** Baseline Characteristic.

| Characteristic | Patient Number<br>(n-57) |
| --- | --- |
| Sex, n (%) |  |
| - Male | 38 (66.7) |
| - Female | 19 (33.3) |
| Age (mean ± SD) | $52.88 \pm 14.04$ |
| HCV antibody, n (%) |  |
| - Reactive | 36 (63.2) |
| - Non-reactive | 21 (36.8) |

#### Concordance Between The Assays

A Total of 57 patients, seven were detected by both GeneXpert HCV VL Fingerstick and GeneXpert HCV Viral Load in conjunction with Cobas Taqman 96. The probability test comparing HCV FL FS, HCV Viral Load, and Cobas Taqman 96 revealed a p-value of <0.001, indicating a statistically significant correlation. Both the HCV FL FS and HCV Viral Load viral load assays demonstrated 100% sensitivity, specificity, negative predictive value (NPV), and positive predictive value (PPV). (Table 2,3)

**Table 2.**
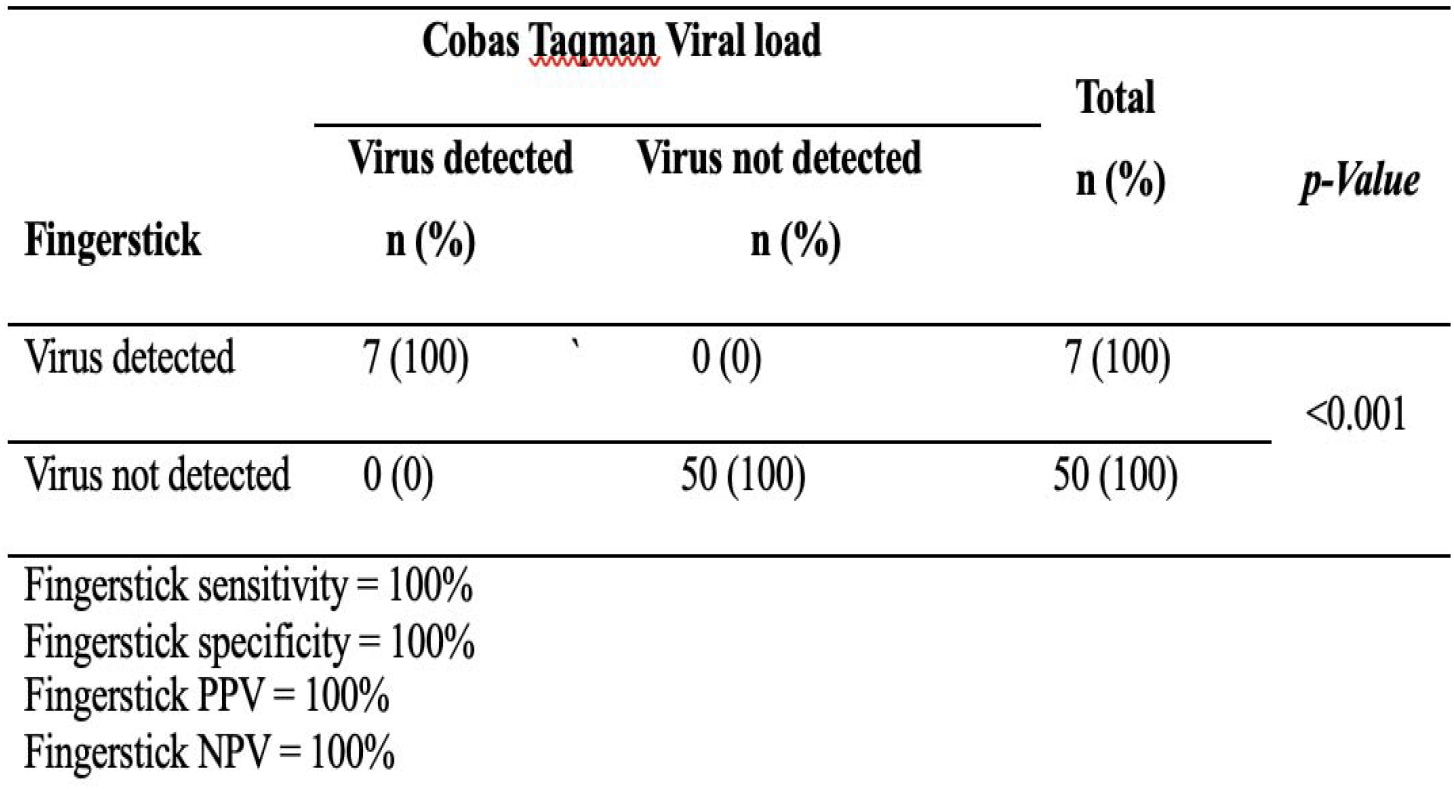
Qualitative results of viral load in Fingerstick and Cobas Taqman 96.

**Table 3.** Qualittive results of viral load in Genexpert and Cobas Taqman 96.

| Cobas Taqman Viral load |  |  |  | *p-Value* |
| --- | --- | --- | --- | --- |
|  | Virus detected | Virus not detected | Total |  |
| Genexpert | n (%) | n (%) | n (%) |  |
| Virus detected | 7 (100) | 0 (0) | 7 (100) | <0.001 |
| Virus not detected | 0 (0) | 50 (100) | 50 (100) |  |
| Genexpert sensitivity = 100% |  |  |  |  |
| Genexpert specificity = 100% |  |  |  |  |
| Genexpert PPV = 100% |  |  |  |  |
| Genexpert NPV = 100% |  |  |  |  |

### Qualitative performance and diagnostic accuracy Xpert® HCV VL FS and Plasma test

#### GeneXpert HCV VL Fingerstick

When compared to the reference test, the GeneXpert HCV FS Viral Load assay using capillary blood showed 100% sensitivity and 100% specificity, with both the positive and negative predictive values also reaching 100%.

#### GeneXpert HCV Viral Load

Compared to the reference test, the GeneXpert HCV VL assay using capillary blood demonstrated 100% sensitivity and 100% specificity, with both the positive and negative predictive values also at 100%.

#### Correlation Between The Assays

Through linear regression analysis of the quantifiable viral loads among 57 participants for comparison of the GeneXpert HCV VL Fingerstick, and GeneXpert HCV Viral Load, RT PCR COBAS TaqMan®, the sensitivity of the the GeneXpert HCV VL Fingerstick, and GeneXpert HCV Viral Load assay quantification in capillary whole blood samples collected by finger stick was 100% and highly significant a strong correlation was observed between the two tests: Fingerstick vs. Cobas Taqman 96 (Fig.1) and GeneXpert Viral Load vs. Cobas Taqman 96 (Fig.1) (R = 0.9569 and R = 0.9602, p = 0.001). Additionally, Figure 3 demonstrated an exceptionally strong correlation between Fingerstick and GeneXpert VL (R = 0.9983, p = 0.001)

**Figure 1.**
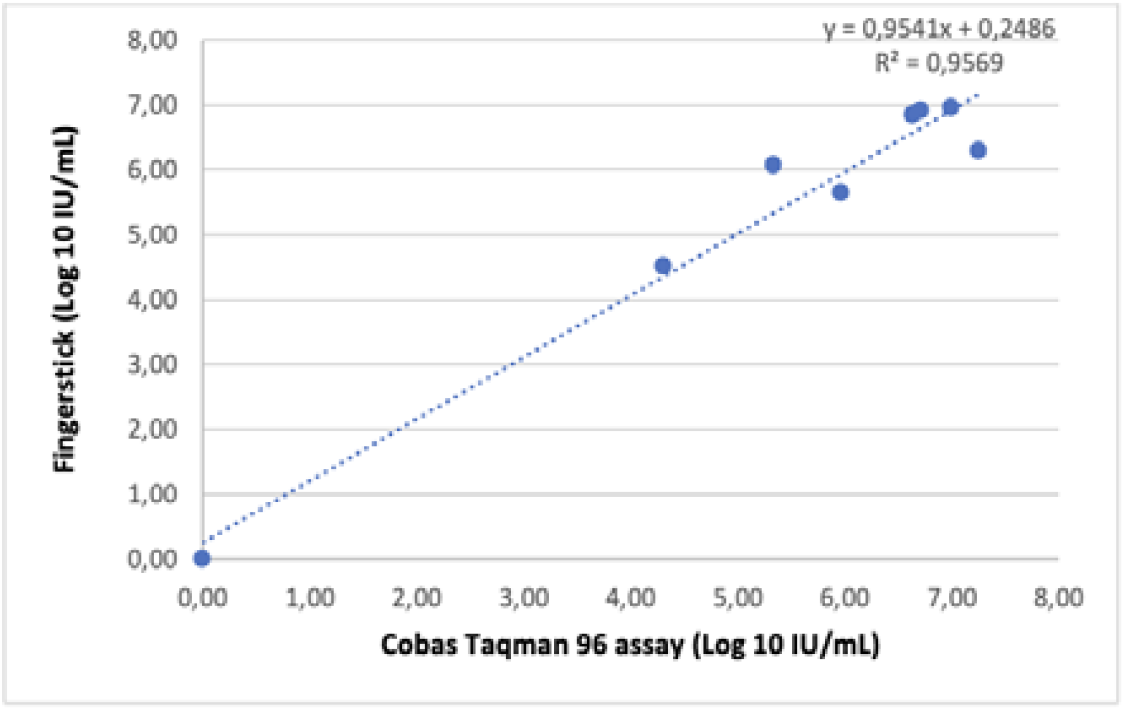
Regression Linear Analysis of Fingerstick vs Cobas Taqman 96.

**Figure 2.**
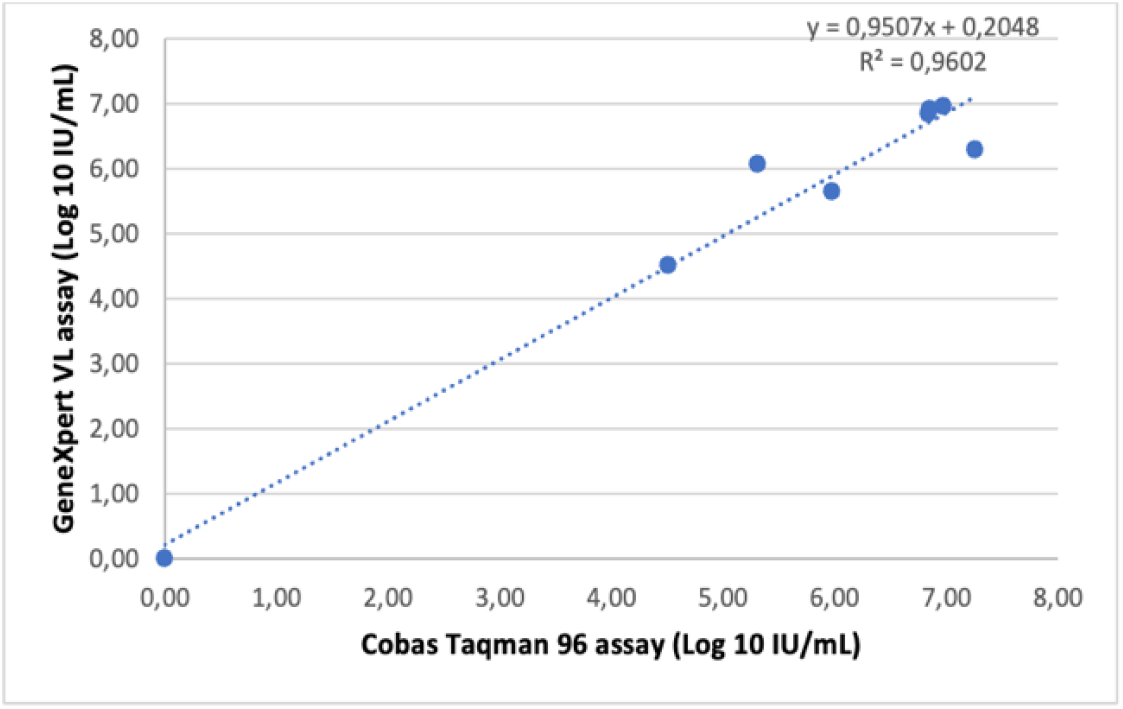
Regression Linear Analysis of GeneXpert VL vs Cobas Taqman 96.

**Figure 3.**
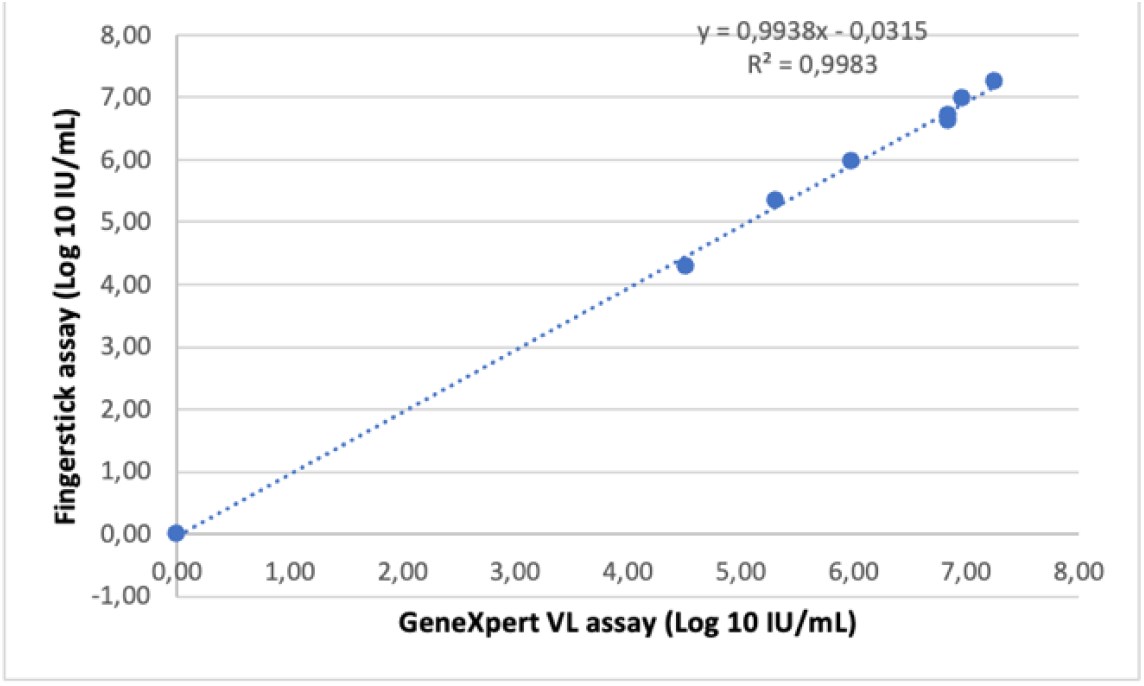
Regression Linear Analysis of Fingerstick VL vs Cobas GeneXpert VL.

## Discussion

The GeneXpert HCV VL FS assay represents a significant advancement over the GeneXpert HCV Viral Load assay. Many participants in the study had previously received hepatitis C treatment and achieved SVR, which resulted in the detection of a substantial number of negative viral results. This highlights the effectiveness of the GeneXpert system in identifying viral load. A study conducted in Tanzania found excellent correlation and concordance in HCV RNA levels between plasma samples and finger-stick samples, demonstrating the clinical utility of this technology. These findings suggest that finger-stick sampling can reliably replace venous blood collection for HCV viral load testing, a crucial step in the cascade of care. The strong correlation (R^2^ values above 0.95) between these methods and Cobas Taqman 96 further supports their clinical reliability. The study also highlighted the potential of the GeneXpert® HCV Viral Load Fingerstick assay in supporting efforts to eradicate HCV in Africa. Given that most individuals with the infection remain undiagnosed, this test could play a vital role in improving detection rates across the continent. (9) (16) A related study by Bregenzer et al. in 2019 has demonstrated that the methodology addresses a significant barrier to HCV treatment particularly for patients with limited venous access and confirmed previous research, establishing that whole-blood testing exhibits sensitivity comparable to venous plasma testing, except in instances where HCV RNA levels are exceedingly low. The performance of the tests was further evaluated across several clinical contexts, including pre-treatment, on-treatment follow-up, end-of-treatment (EOT), sustained virological response (SVR), reinfection screening, and the differentiation between spontaneous clearance and chronic infection. From 111 patients, the correlation analysis for quantifiable results (log IU/ml) between the GeneXpert® tests and the Cobas® test demonstrated a strong relationship, with correlation coefficients (R^2^) of 0.9165 for the GeneXpert HCV VL test and 0.9899 for the GeneXpert HCV VL FS test. Both tests exhibited a high degree of concordance with the reference RT PCR Cobas Taqman test, further supporting their clinical reliability. Accuracy evaluation under various conditions showed that both tests maintained high sensitivity and specificity, particularly for pre-treatment baseline and sustained virological response (SVR) monitoring, with sensitivity rates of 97.0% for GeneXpert HCV VL and 100% for GeneXpert HCV VL FS test. nevertheless, some discordant results, including false positives and false negatives, were observed, primarily in cases where HCV RNA levels were near the limit of quantification (LOQ), which was most common during direct-acting antiviral (DAA) treatment.(17) In a similar study conducted by Grebely et al. has demonstrated the GeneXpert HCV Viral Load POC assay using both venepuncture-collected and finger-stick blood samples. The researchers reported that the finger-stick samples had a sensitivity of 97.7% and a specificity of 99.1% for HCV RNA quantification, suggesting that this approach may be a viable option for HCV diagnosis in resource-limited settings.(18) The virus is one of the main contributors to chronic liver disease., significantly contributing to the burden of cirrhosis, liver failure, and hepatocellular carcinoma. Accurate viral load measurements are essential for monitoring treatment response and guiding clinical decision-making. (19) POCT for HCV-RNA has become a valuable tool for simplifying the diagnosis of active HCV infection and enabling immediate treatment to begin, marking an important breakthrought in the diagnosis and management of hepatitis. (20,21) In Italy, Calvaruso et al. in 2019 for the first time at Week 4, 39/56 (69.6%) and 42/56 (75%) patients had undetectable HCV□RNA by GeneXpert HVL Finger Stick and RT□PCR test, respectively. The ere six patients showed undetectable HCV RNA by RT□PCR, even though levels <10 IU/mL using “The GXHVL” test, 5 patients had undetectable HCV RNA using the Gen Xpert HVL Finger Stick, but <15IU/mL by RT-PCR. Both test showed undectectbale HCV RNA in all 56 patients (100%) at the end of treatment (EOT). At SVR 12, both tests identified a single case of HCV relaps in the cohort, with 100% a agreement rate. This also demonstrated the feasibility of using POCT’s for detecting HCV RNA and evaluating viral respone to antiviral treatment. The HCV RNA testing using fingerstick blood sample and analyzed with the GeneXpert POC provided results that were fully comparable to the laboratory-based tests, even in the setting of managing HCV antiviral treatment (23) Although the findings are encouraging, the study has certain limitations, particularly the relatively small sample size. Future studies with larger cohorts are needed to validate these results and evaluate the long-term effectiveness of these alternative testing methods in clinical settings.

## Conclusion

Few studies have reported on the sensitivity, specificity, and rapid results of the newly developed GeneXpert Fingerstick HCV Viral Load assay for HCV RNA quantification using capillary blood collected via fingerstick, with results available in just one hour. When compared to the RT-PCR method, this study showed a strong correlation between the qualitative and quantitative detection of the virus. The availability of similarly accurate POCTs for other viral hepatitis markers, like HBV or HAV, would play an important role in the comprehensive control of viral hepatitis. In summary, this study demonstrates that finger-stick blood testing promotes the continued evolution of Hepatitis C diagnosis and treatment through the implementation of a rapid, accessible and less invasive testing method, making it a game changer in providing access to a disease typically associated with under-served populations and the opportunity for providers to reach into these gap groups to facilitate better care and outcomes. If implemented, finger-stick-based HCV viral load monitoring would be a positive step towards achieving the World Health Organization’s 2030 target of eliminating HCV worldwide.

## Data Availability

All relevant data are within the manuscript and its Supporting Information files.

